# Incremental Value of Bi-Ventricular Ejection Fraction (BiVEF) Phenotyping for the Discrimination of Heart Failure Symptoms and Clinical Outcomes: A Cardiovascular Magnetic Resonance Study of 9,437 Patients

**DOI:** 10.1101/2025.04.18.25326093

**Authors:** Edward Nabet, Dina Labib, Steven Dykstra, Shahidul Islam, Jacqueline Flewitt, Sandra Rivest, Wendy Drewes, Shant Manoushagian, Carmen P. Lydell, Andrew G. Howarth, Richard Chazal, Malissa Wood, Katherine Lietz, Kevin Marzo, Juan Lopez-Mattei, Nowell Fine, James A. White, Juan Gaztanaga

**Author notes:** These authors contributed equally to this work and share first authorship (Edward Nabet, Dina Labib). These authors contributed equally to this work and share last authorship (James A. White, Juan Gaztanaga).

## Abstract

**Background:** Left ventricular ejection fraction (LVEF) continues to be employed as the principle phenotypic marker for the classification, prognostication, and management of cardiovascular disease.

However, expanding evidence identifies similarly important roles for right ventricular ejection fraction (RVEF) across multiple referral cohorts, raising the consideration of bi-ventricular ejection fraction (BiVEF) based phenotyping in broader clinical practice. We assessed the value of cardiovascular magnetic resonance (CMR)-based BiVEF phenotyping versus conventional LVEF-only phenotyping for the prediction of NYHA functional class and future heart failure (HF) outcomes.

**Methods:** 9,437 consecutively enrolled adult patients clinically referred for CMR were evaluated for NYHA class ≥II at time of imaging and a future composite outcome of HF hospitalization, HF death, and need for cardiac transplantation or LV assist device.

**Results:** Median age was 57 years (Q1, Q3 44-66, 62% male). Across all LVEF strata, RVEF<45% was independently associated with NYHA ≥II after comprehensive adjustment for baseline clinical and imaging characteristics. Respective adjusted odds ratios for RVEF <45% versus ≥45% were 2.30 (1.79-2.97), 1.58 (1.13-2.20), and 2.01 (1.44-2.79) for LVEF <40%, 40-50, and >50% categories (p<0.001, =0.007, and <0.001; respectively). Over a median follow-up of 3.9 years, 766 patients (8%) experienced the HF outcome. In a multivariable Fine-Gray model, the respective adjusted HR_sub_ for LVEF <40% and 40-50% were 2.13 (1.64-2.77) and 1.70 (1.33-2.17); p<0.001) relative to LVEF >50%. In this model, RVEF <45% was associated with 1.52 (1.25-1.86) greater hazard for future HF outcome versus RVEF ≥45% (p<0.001).

**Conclusions:** RV contractile health is independently associated with HF symptoms and identifies patients at elevated risk of future HF outcomes. Incremental prognostic value from BiVEF phenotyping is delivered versus LVEF-only phenotyping.

## INTRODUCTION

Cardiovascular diseases are the leading cause of morbidity and mortality worldwide, accounting for ∼18.6 million deaths in 2019.^1,2^ Related healthcare costs in the United States were estimated at 393 billion in 2020 with a projected increase to 1,490 billion by 2050.^3^ Left ventricular (LV) ejection fraction (LVEF) has been widely adopted as the primary phenotypic marker of eligibility for clinical trials and cardiovascular therapeutics over the past several decades, a practice justified by its clinical availability, rational physiologic representation of cardiomyopathy severity, and demonstration of predictive value for major cardiovascular outcomes (MACE)^4–6^. However, recognition that LVEF phenotypes alone do not explain heart failure (HF) symptoms or future HF outcomes, particularly in patients with preserved LVEF^7,8^, has generated substantial interest surrounding the role of right ventricular (RV) health in the prediction of outcomes across focused^9–16^ and broad^3,17,18^ referral populations. Additionally, while some studies have explored the associations of RV dysfunction with impaired exercise capacity as assessed by cardio-pulmonary exercise testing^19–21^, data surrounding its direct association with HF symptoms assessed by the New York Heart Association (NYHA) tool are sparse^22^. The latter is an objective index of functional capacity routinely captured during the assessment of patients with cardiac disease, has established prognostic value, and is widely used to guide therapeutic decisions.^22–24^

RV contractile health can be assessed by trans-thoracic echocardiography (TTE) using the pragmatic surrogates of tricuspid annular plane systolic excursion (TAPSE) and fractional area change; less commonly using speckle tracking echocardiography or 3D echocardiography.^25, 26^ While 3D echocardiography has shown robust agreement with cardiovascular magnetic resonance (CMR) imaging^27^, the latter remains the reference standard for RV phenotyping based upon the accurate quantification of RV volumes^28,29^ with incremental RV-relevant markers of myocardial remodeling, such as RV insertion (RVI) site fibrosis^30^. By this technique, a prior study of 7,131 patients with cardiovascular disease reported by our group showed incremental value from RV ejection fraction (RVEF) for the prediction of composite MACE outcomes and a focused composite HF outcome, independent of LVEF.^18^

Justified by our prior study, and leveraging an expanded cohort of 9,437 unique patients, this study aimed to assess the value of implementing CMR-based bi-ventricular ejection fraction (BiVEF) phenotyping versus conventional LVEF-only phenotyping in patients with known or suspected cardiovascular disease. Phenotype classification schemas were compared with respect to their independent associations with New York Heart Association (NYHA) functional class and future heart failure outcomes.

## METHODS

### Study Design and Patient Population

This was an a-priori sub-study of the Cardiovascular Imaging Registry of Calgary (CIROC), a prospectively recruiting observational cohort study of patients clinically referred for CMR imaging in Southern Alberta, Canada, launched in January 2015. As previously described^18^, consenting patients complete tablet-based health questionnaires for the collection of core demographics, socio-demographics and habits, cardiovascular symptoms, and health-related quality of life prior to cardiac imaging test performance (intakeDI^TM^, Cohesic Inc., Calgary, AB). Cardiac phenotypes are captured by standardized reporting (cardioDI^TM^, Cohesic Inc., Calgary, AB), followed by iterative migration and secure matching of administrative health data from local institutional data warehouses, inclusive of laboratory, pharmacy, and ICD-10-CA (Canadian version of ICD-10)-coded clinical variables.

On February 28th, 2022, 10,741 Registry-enrolled patients had completed index CMR assessments, baseline health questionnaires, and a minimum of 6-months clinical follow-up. For the current analysis, we included patients ≥18 years old without congenital disease, recognizing unique structural anatomy and/or loading conditions on the RV. Specifically, patients with tetralogy of Fallot (TOF), tricuspid valve atresia, ventricular septal defect, atrial septal defect, anomalous pulmonary vein drainage, double outlet RV, hypoplastic heart syndromes, and Ebstein anomaly were excluded.

### Patient Health Questionnaires and Cardiovascular Risk Factors

Standardized health questionnaires were deployed to capture patient-reported ethnicity, socio-demographics, smoking habits, and health-related quality of life using the EQ-5D tool, as well as cardiovascular symptomatology at time of CMR imaging. The latter included patient-reported shortness of breath using the NYHA classification as well as chest pain using the Canadian Cardiovascular Society (CCS) Angina Score. Definitions for cardiovascular risk factors and comorbidities were established using a combination of patient questionnaires and administrative health data, as described in **Supplemental Methods**.

### CMR Imaging and Analysis Protocol

CMR imaging was performed using 3 Tesla clinical scanners (Prisma or Skyra, Siemens Healthcare, Erlangen, Germany). Standardized CMR imaging protocols were used, inclusive of cine imaging using a steady-state free precession pulse sequence in sequential short-axis views from above the pulmonary valve to beyond the cardiac apex. Long-axis imaging was performed in the 2-, 3-and 4-chamber views. Late gadolinium enhancement (LGE) imaging was performed 10 minutes following gadolinium contrast administration (0.15–0.2 mmol/kg; Gadovist; Bayer, Inc) using a standard inversion recovery gradient echo pulse sequence in matched views to cine imaging.

Quantitative image analysis was performed using commercially available software (cvi**42**; Circle Cardiovascular Imaging Inc, Calgary, Canada) applying standardized operational procedures adherent to published Society of Cardiovascular Magnetic Resonance recommendations^31^. Details on CMR analysis techniques are included in **Supplemental Methods**.

### Clinical outcomes

This study explored two primary outcomes. The first was patient-reported NYHA class at time of baseline CMR imaging. The second was the future occurrence of a composite of heart failure outcome, inclusive of heart failure mortality, heart failure hospitalization, cardiac transplantation, or left ventricular assist device (LVAD) implantation. Cause of death was adjudicated by electronic medical chart review utilizing definitions from the 2017 Cardiovascular and Stroke Endpoint Definitions for Clinical Trials consensus report^32^. Remaining outcomes were based on ICD-10-CA and procedural Canadian Classification of Health Interventions (CCI) coding. A full list of codes used for the outcomes is shown in **Supplemental Methods**.

### Statistical Analysis

Continuous variables were expressed as mean (standard deviation [SD]) or median (Q1, Q3); categorical variables as counts (percentage). Comparisons of low versus high RVEF categories for baseline variables were performed using two-sample t-test/Mann-Whitney test for continuous variables or Chi-square/Fisher exact test for categorical variables.

A multivariable logistic regression model was constructed to test the association of LV and RVEF with NYHA ≥II, with results reported as odds ratios (OR) and 95% confidence intervals (CI).

Time to the composite heart failure outcome were calculated as the time from baseline CMR imaging until the first outcome occurrence. Patients who did not develop the outcome were considered censored at the time of last follow-up or occurrence of non-heart failure death.

Cumulative incidence functions (CIFs)^33^ were used to estimate the incidence of the composite outcome, using Gray’s test^34^ to compare CIFs between RVEF categories. Multivariable competing risk Fine-Gray models^35^ were constructed to test associations of LVEF and RVEF with the heart failure outcome (R packages ‘cmprsk’), considering non-heart failure death as a competing event. Results were reported as subdistribution hazard ratios (HR_sub_) and 95% CI.

For each of the multivariable logistic regression and Fine-Gray models, LVEF and RVEF were adjusted for CMR referral indication and pre-specified baseline non-imaging and imaging variables with known clinical relevance. Only variables with an overall missingness rate <20% were considered. LVEF and RVEF were modelled both as continuous variables and categorical variables using the pre-specified categories of <40%, 40-50%, and >50% for LVEF and <45% and ≥45% for RVEF. Choice of the LVEF categories was based on convention as recommended by ACC/AHA/HFSA guideline for the management of heart failure^36^, whereas choice of RVEF categories was based on the cut off value adopted by prior CMR studies^10,12,14,37^ as well as being identified as a clinical threshold to define abnormal RV function by 3D echocardiography^251^.

Missing covariates in the multivariable models were imputed using the *aregImpute* algorithm^38^, with details provided in **Supplemental Methods**.

Analyses were conducted using R, version 4.4.1, with a two-tailed P<0.05 indicating statistical significance.

## RESULTS

### Baseline characteristics

A total of 9,437 patients met the study inclusion criteria and were used for analysis. Baseline characteristics are shown in **Table 1**. Median age was 57 (Q1, Q3 44, 66) years; 62% being male. CAD was the most common referral indication. Distribution of patients across LVEF categories was 1,392 (15%), 1,201 (13%), and 6,844 patients (73%) for LVEF <40%, 40-50%, and >50%, respectively. A total of 1,099 patients (12%) had an RVEF <45%. LGE imaging was performed in 7,773 patients (82%), with 3,289 (42%) having LGE patterns other than RVI site fibrosis.

**Table 1.**
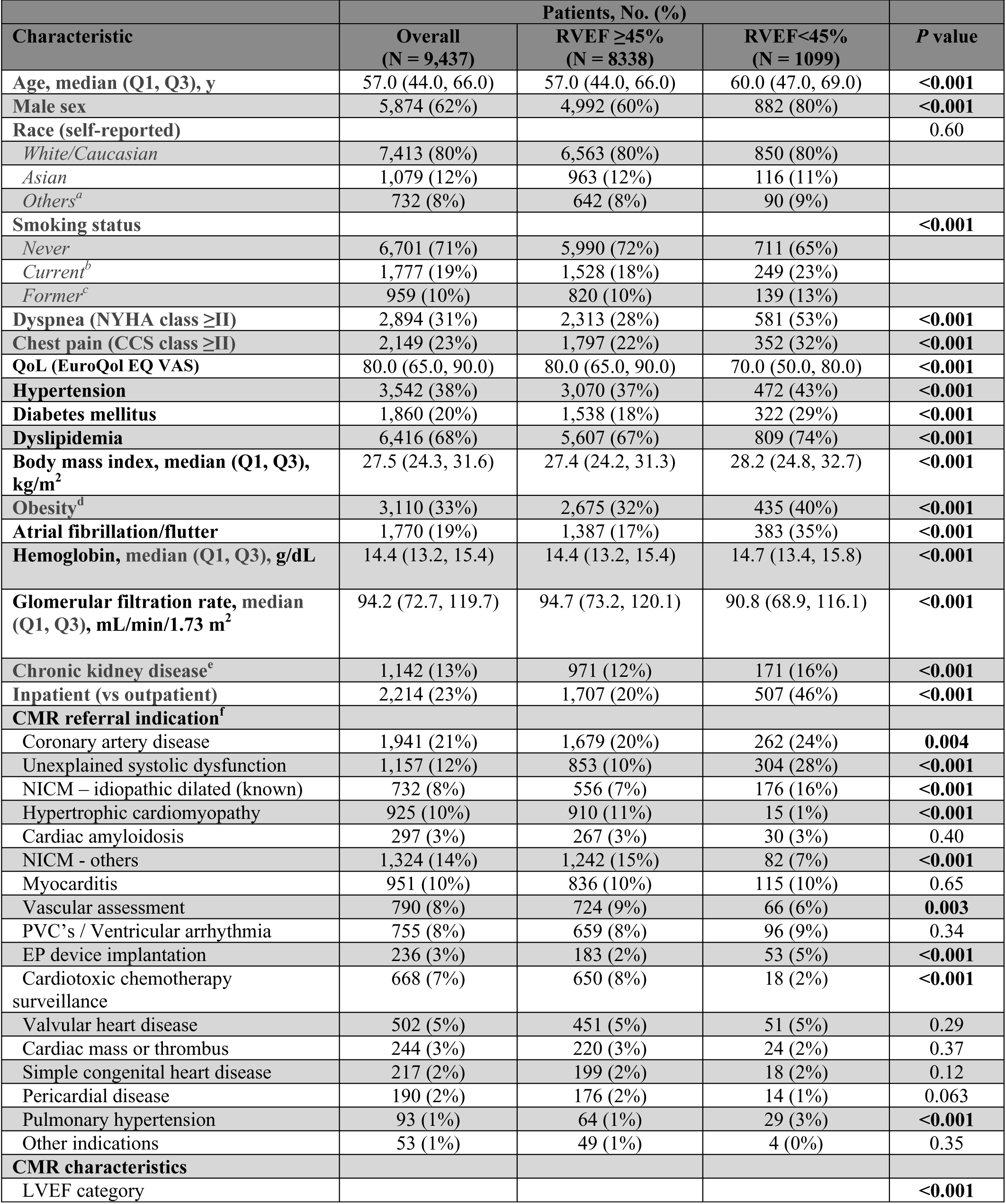
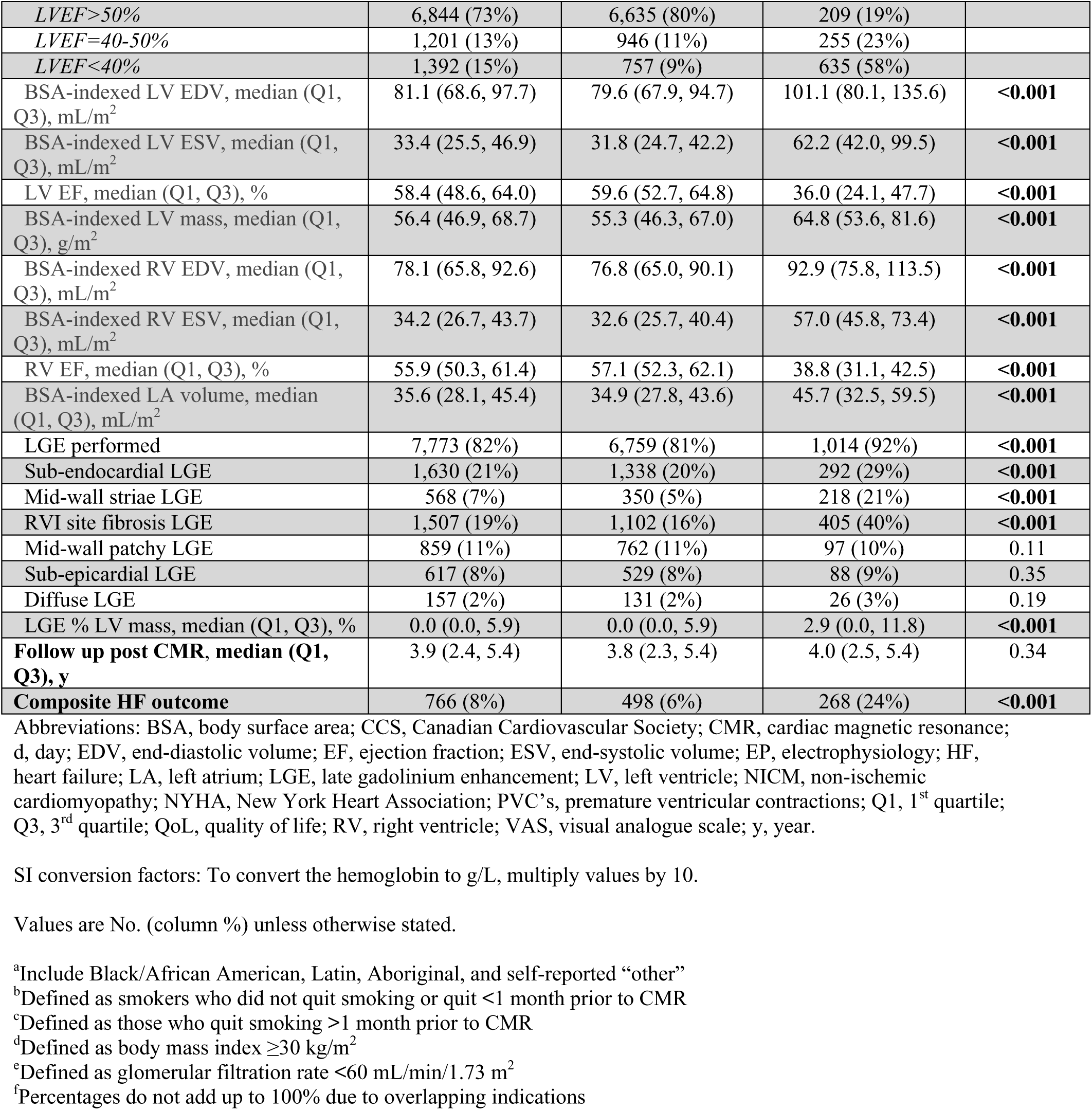
Baseline characteristics of the study population, stratified according to categories of right ventricular ejection fraction (RVEF)

As shown in **Table 1**, compared to patients with an RVEF ≥45%, patients with an RVEF <45% showed significant differences in baseline clinical characteristics, inclusive of a higher proportion of males and smokers, as well as an increased prevalence of diabetes, hypertension, obesity, and atrial fibrillation/flutter. Patients with an RVEF <45% also showed worse CMR - based cardiac phenotypes in the form of dilated cardiac chambers, worse LVEF, and a higher fibrosis/scar burden.

### Associations of BiVEF Phenotypes with NYHA Functional Class

In a multivariable logistic regression model, both LVEF and RVEF were each significantly associated with NYHA class ≥II, following adjustment for baseline characteristics (**Supplemental eFigure 1**). Respective adjusted OR for NYHA class ≥II (95% CI) were 0.77 (0.71-0.83) and 0.75 (0.69-0.81) per 5% increase in LVEF and RVEF (p<0.001 for each). A significant interaction was observed between LVEF and RVEF (p< 0.001 for the interaction term). To simplify explanation of this interaction effect, continuous LVEF and RVEF values were replaced by categorized variables with the calculation of the odds for RVEF <45% versus ≥45% in each LVEF category. In this model, compared to patients with preserved LVEF >50%, those with LVEF of 40-50% and <40% experienced a 1.13-fold (0.94-1.35; p=0.2) and 1.36-fold (1.09-1.69; p=0.006) higher odds of NYHA class ≥II status, following adjustment for baseline characteristics (**Figure 1**). Additionally, RVEF <45% showed higher odds of NYHA class ≥II status vs RVEF ≥45% within each LVEF category, demonstrating respective odds of 2.30 (1.79-2.97), 1.58 (1.13-2.20), and 2.01 (1.44-2.79) for patient with LVEF <40%, 40-50%, and >50% (respective p<0.001, =0.007, and <0.001; **Figure 2**). Overall model AUC was 0.79 (0.78-0.80) for both models with continuous and categorized values. Compared to the multivariable models including only LVEF, the addition of RVEF demonstrated incremental value using both continuous and categorized values, with a respective increase in the Chi-square value of 69.7 and 64.1 (p<0.001 for each model using the likelihood ratio test).

**Figure 1.**
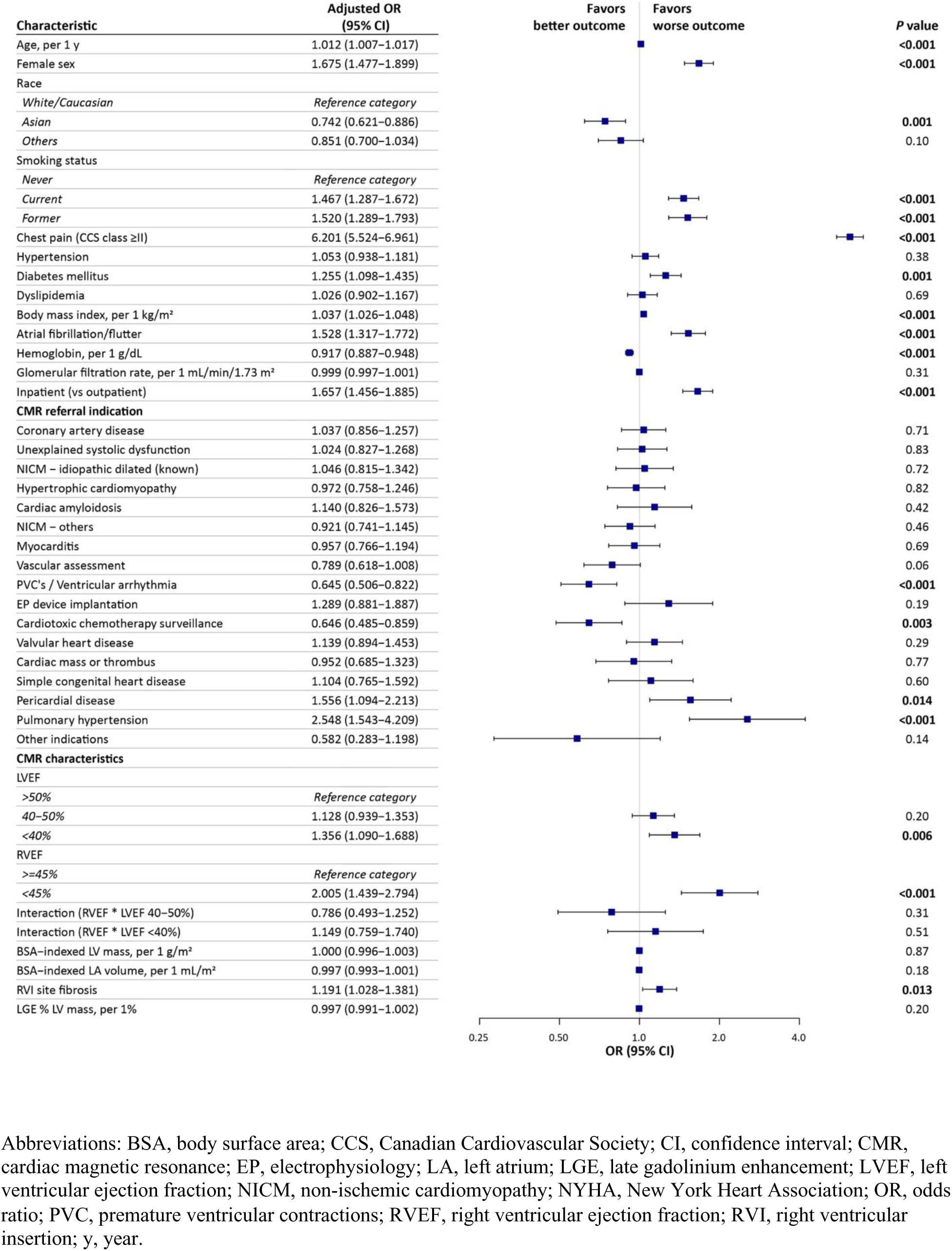
Forest plot of the results of the multivariable logistic regression model for associations of left ventricular and right ventricular ejection fraction, modelled as categorical variables, with NYHA functional class ≥II at time of CMR imaging.

**Figure 2.**
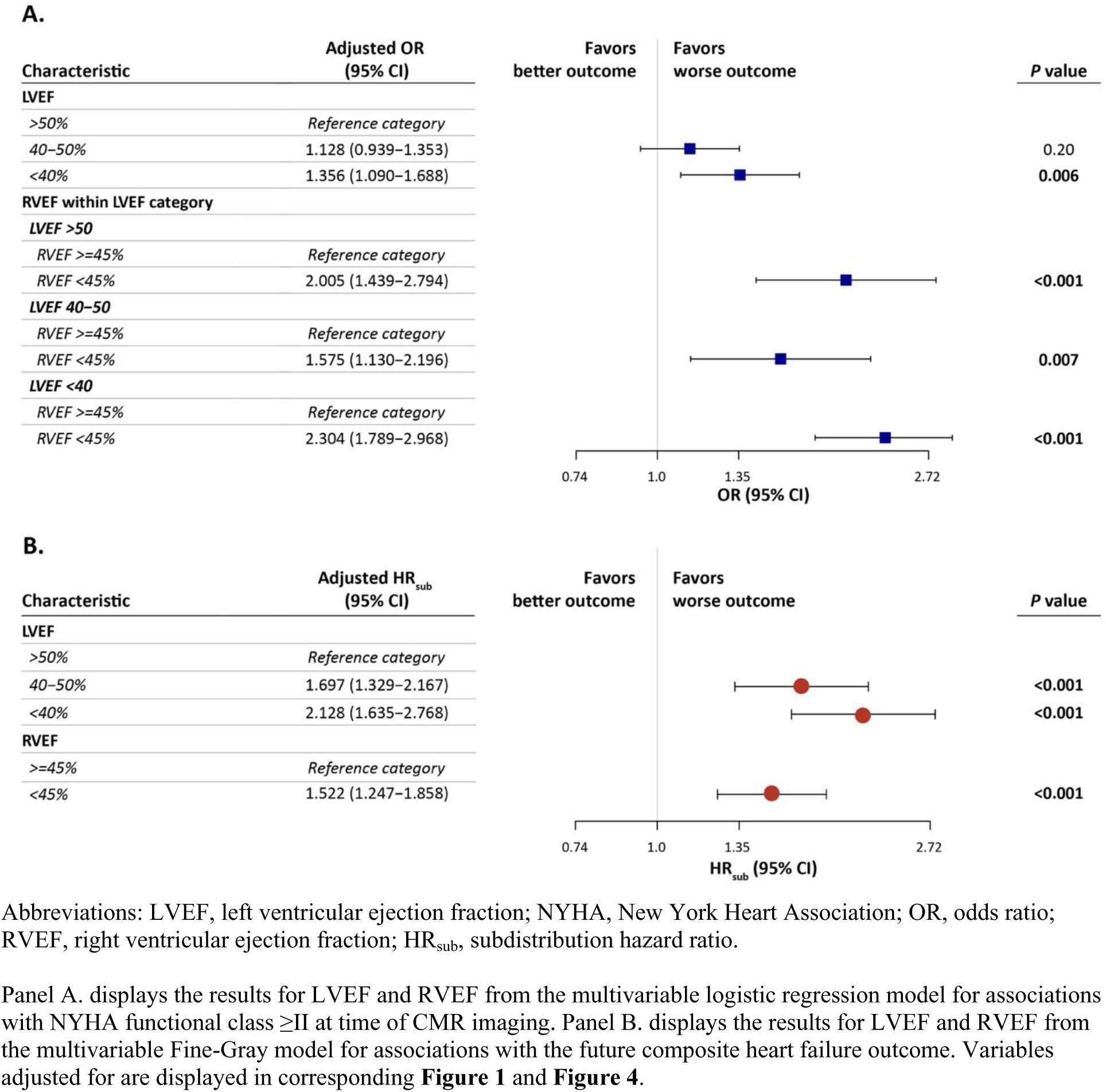
Forest plot for adjusted associations of biventricular ejection fraction (BiVEF) phenotype, modelled as categorical variables, with NYHA functional class ≥II and the composite HF outcome

### BiVEF versus LVEF Phenotypes for the Prediction of the HF Composite Outcome

At a median follow-up of 3.9 years, 766 patients (8%) had experienced an adverse HF outcome. Of these, the first registered outcome was HF hospitalization in 755 patients (99%), HF death in 8 patients (1%), and LVAD placement in 3 patients (0.4%).

Patients with RVEF <45% showed a higher unadjusted 1-year cumulative incidence of the composite HF outcome versus RVEF ≥45% in the overall study population (p<0.001; **Figure 3**). This was also true within each LVEF category (p<0.05 for each). Similar trends were noted for the 5-year cumulative incidence.

**Figure 3.**
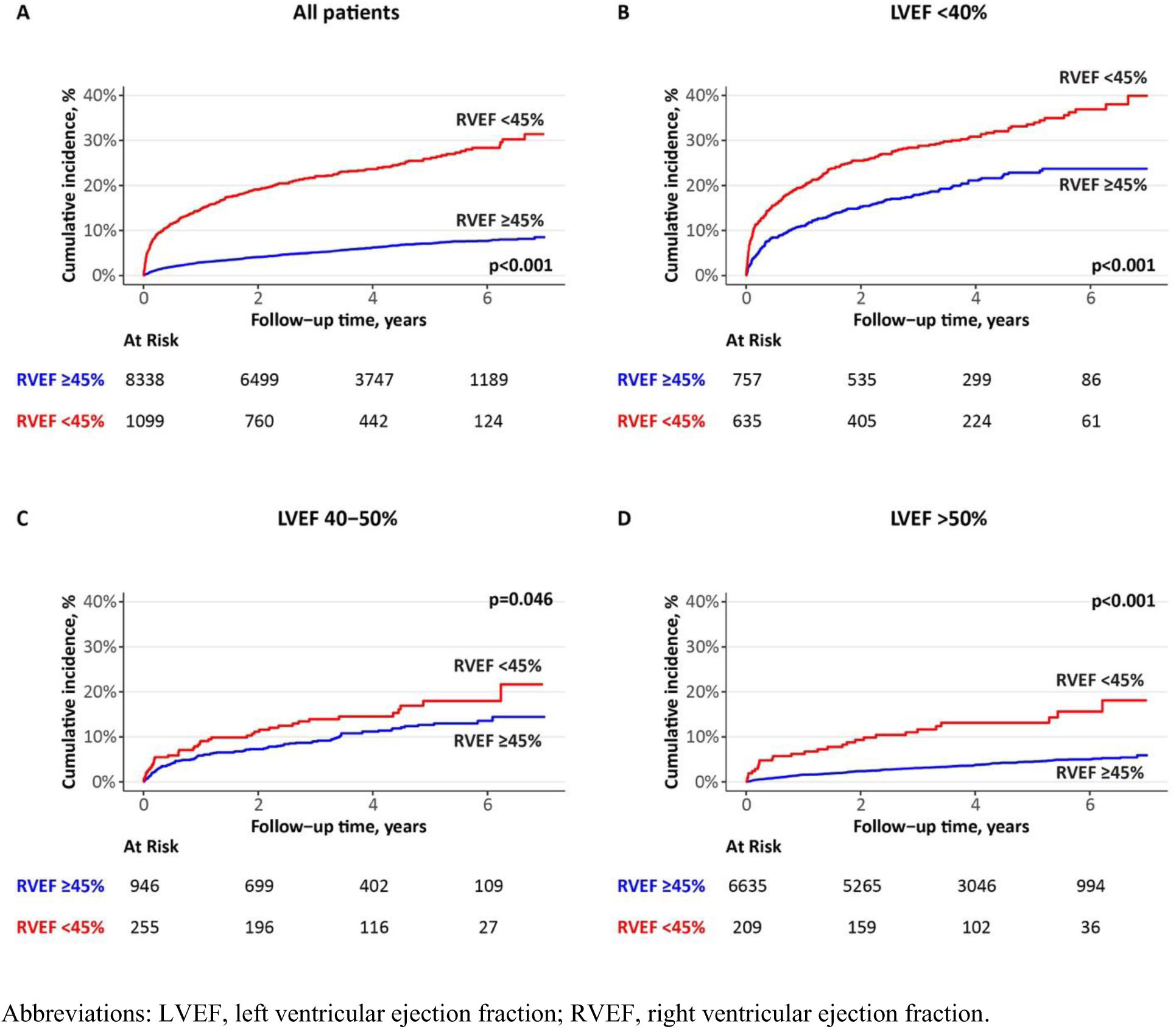
Cumulative incidence plots of the composite outcome of heart failure for the categories of RVEF

Adjusting for the same variables used in the prior logistic regression models, in addition to NYHA ≥II functional class, both LVEF and RVEF on a continuous scale were significantly associated with the composite outcome (respective adjusted HR_sub_ 0.909 (0.871-0.948); p<0.001 and 0.930 (0.890-0.972) per 5% increase in EF; p=0.001; **Supplemental eFigure 2**). Contrary to the logistic regression models, there was no significant interaction between LVEF and RVEF (p-value = 0.12 for the interaction term), denoting that the effect of RVEF is not modified by LVEF and vice versa. Accordingly, the multivariable model was repeated with categorized LVEF and RVEF without including an interaction term and findings reported for each variable separately (**Figure 4**). In this model, the respective adjusted HR_sub_ for LVEF 40-50% and <40% were 1.70 (1.33-2.17) and 2.13 (1.64-2.77) relative to LVEF >50% (p<0.001 for both). The corresponding adjusted HR_sub_ for RVEF <45% was 1.52 (1.25-1.86) versus RVEF ≥45% (p<0.001). Overall model concordance index at the median follow-up of 3.9 years was 0.88 (0.86-0.89) for both models with continuous and categorized values. Similar to the logistic regression models, the addition of RVEF to multivariable models to an LVEF-only model demonstrated incremental value for using both continuous and categorized EF values, with a respective increase in the Chi-square value of 12 and 20 (p=0.001 and <0.001).

**Figure 4.**
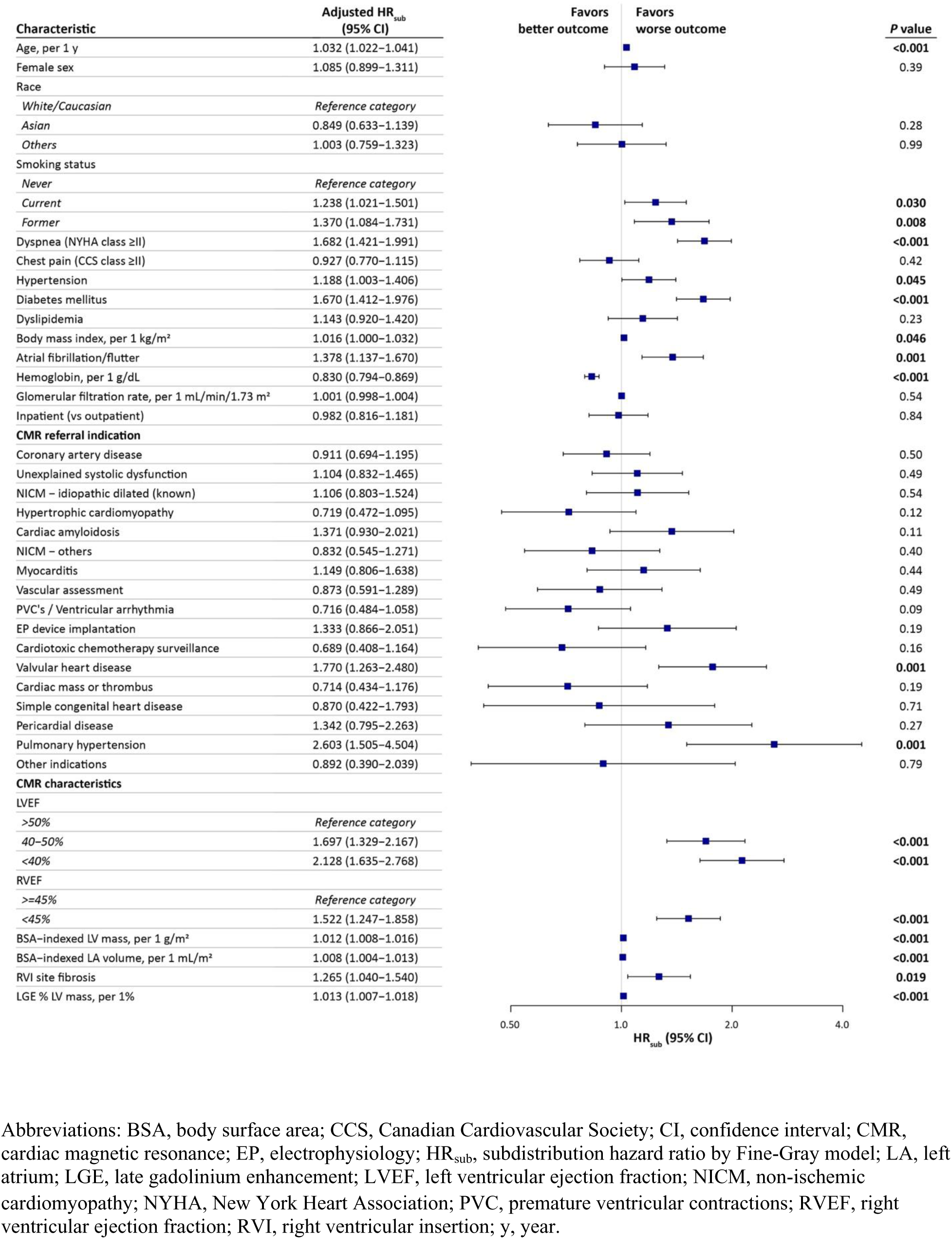
Forest plot of the results of the multivariable Fine-Gray model for associations of left ventricular and right ventricular ejection fraction, modelled as categorical variables, with the future composite heart failure outcome.

### Associations of RVI site fibrosis with RVEF, NYHA functional class, and the HF outcome

Patients with RVEF <45% demonstrated a higher prevalence of RVI site fibrosis compared to RVEF ≥45% (40% versus 16%; p<0.001; **Table 1**). RVI site fibrosis was independently associated with a higher odds of NYHA ≥II functional class (adjusted OR 1.19 [1.03-1.38; p=0.013; **Figure 1**) as well as future development of the composite HF outcome (adjusted HR_sub_ 1.27 [1.04-1.54]; p=0.019; **Figure 4**) in the models with categorized LVEF and RVEF values.

## DISCUSSION

This is the largest reported investigational study to date evaluating the incremental value of RVEF to LVEF alone for explaining patient HF symptomatology and future composite HF outcomes across a broad cardiovascular disease referral population. Using the reference standard of CMR-based quantification, an RVEF <45% was shown to provide incremental utility for the prediction of NYHA function class ≥II across all conventional LVEF strata, following comprehensive adjustment for baseline clinical characteristics and CMR clinical referral indication. Additionally, we demonstrated incremental prognostic value of RVEF beyond LVEF with respect to future HF outcomes irrespective of LVEF range. RVI site fibrosis was more prevalent among patients with RV dysfunction, showing independent associations with both functional capacity and HF outcomes.

To the best of our knowledge, our study is the first to report on direct associations of RVEF and NYHA functional class within a large, broad cardiac disease referral cohort, benefiting from comprehensive adjustment for baseline clinical and imaging variables. A prior small sized study of 100 late survivors following TOF surgical repair reported on the independent association of CMR-measured RVEF with NYHA class ≥III, reporting an adjusted OR 2.4 (95% CI, 1.2-4.7) per 10% decrease in RVEF.^39^ Another study of 565 patients with normal LVEF, reported on the univariable associations of impaired systolic and diastolic RV strain, measured by echocardiography, with NYHA functional status.^40^ Finally, a third study explored univariable associations of radionuclide angiography-based RVEF with exercise capacity measured by cardio-pulmonary exercise testing in 25 patients of ischemic and non-ischemic cardiomyopathy.^19^

Several mechanisms can justify the additive contribution of RVEF to HF symptoms (NYHA functional class) incremental to LVEF. RV dysfunction can exist despite normal LVEF, this demonstrated by 19% of our patients with LVEF >50% having an RVEF <45%. This has been similarly supported in a previous study of heart failure with normal LVEF where the prevalence of systolic and diastolic RV dysfunction was 75% and 48% based on several conventional and strain based echocardiographic markers.^40^ RV systolic dysfunction has been postulated to produce HF symptoms even with normal LVEF due to low stroke volume resulting in reduced perfusion of both respiratory and peripheral muscles.^41, 42^ Diastolic RV dysfunction can also contribute to HF symptoms through renal venous congestion leading to fluid retention and pulmonary congestion.^43^

RV health is increasingly recognized to be an important determinant of major cardiovascular outcomes.^3,9,10, 11, 13–15,16,18^ While RVEF is commonly acknowledged as a relevant clinical variable in patients with advanced pulmonary disease or right-sided valvular disease, the engagement of RVEF (or validated surrogates of RVEF) as a core phenotypic marker (similar to LVEF) has not been embraced by clinical practice. This must now be recognized to be in contrast to broadening recognition for RV contractile health as a critical predictor of cardiovascular outcomes^22^ in ischemic cardiomyopathy^,44, 45^, non-ischemic cardiomyopathies^10,12^, acute myocarditis^46^, valvular heart disease^47, 48^, and HF with normal LVEF populations^11,40^.

Across these states, numerous pathophysiologic mechanisms may be invoked, inclusive of primary RV myocardial disease, transmissive hemodynamic force via the pulmonary vasculature, and inter-ventricular interactions through shared septal myocardium.^49, 50^ Collectively, these factors contribute to reductions in RVEF that have now been consistently demonstrated to predict future outcomes in disease-specific referral cohorts.^10,13–16^

Despite disease-specific cohort studies demonstrating prognostic value for RVEF, studies justifying value for routine phenotyping using this marker in broad clinical practice are limited. Our team previously reported on 7,131 patients identifying that RVEF <40% by CMR was associated with a 3.1-fold risk of MACE following comprehensive adjustment for baseline demographics, comorbidities, and other non-RV CMR phenotypic markers^183^. In this study each 10% drop in RVEF was associated with a 1.3-fold increased risk of the secondary composite HF outcome. Building on these seminal findings, the current study was designed to examine the value of routine BiVEF phenotyping across clinical practice to better characterize the cardiovascular phenotype with respect to both HF-related morbidity and prognosis. Based on our study findings, the pragmatic stratification of patients based on RVEF <45% and ≥45% was additive to conventional LVEF strata, allowing for significantly improved discrimination of functional impairment and patients at risk of HF-related outcomes.

### Limitations

We acknowledge several important limitations. This is a single center study and therefore external validation is warranted. Inherent to the chosen study design, leveraging the reference standard imaging technique of CMR, modality referral bias must be considered. This may influence the distributions of disease encountered relative to other imaging services. To address this, the referral etiology was routinely captured for all subjects and considered in multivariable models, mitigating this concern for generalization to other referral cohorts. However, it is recognized that clinical translation to other imaging modalities requires independent validation using modality-specific surrogates of RV function. In this study, the assessment of RV health was focused on RVEF and RVI site fibrosis. Assessment of advanced markers inclusive of RV free wall fibrosis and strain is therefore of future interest. Non-invasive or invasive estimates of pulmonary arterial pressures were not captured in this study, thus precluding adjustment for these markers in our models. Finally, we did not adjust for serum biomarkers of tissue injury or systemic inflammation as only clinically ordered laboratory testing was available.

## CONCLUSIONS

This is the largest study to date evaluating the incremental value of RV contractile function across conventional LVEF-defined phenotypes in broad clinical practice. We identified BiVEF phenotyping using the pragmatic approach of stratifying patients according to the presence of a RVEF<45% within each conventional LVEF strata, significantly improves the identification of patients experiencing functional limitation and those at risk of future HF-related outcomes.

These findings support a migration towards biventricular disease phenotyping in broad clinical practice.

## ACKOWLEDGEMENTS

## Data Availability

Data is available upon request and approval and a data sharing agreement.

